# HIGHLY CITED ARTICLES IN EVOLUTIONARY PSYCHIATRY: ASSESSMENT WITH A QUALITY AND ERROR RATING SCALE

**DOI:** 10.1101/2024.07.21.24310766

**Authors:** Chad Beyer, Chanel Robinson, Dan J. Stein

**Affiliations:** Resident, Department of Psychiatry, University of Cape Town, Cape Town, South Africa, 7701; SAMRC Unit on Risk & Resilience in Mental Disorders, Department of Psychiatry & Neuroscience Institute, University of Cape Town, Cape Town, South Africa, 7701

**Keywords:** Evolutionary psychiatry, systematic review, citation analysis, article quality, common errors, rating scale

## Abstract

**Introduction:** Evolutionary psychiatry is a rapidly growing field that emphasizes the value of evolutionary explanations for traits that make individuals vulnerable to mental disorders. Some articles that apply evolutionary theory to psychiatric disorders make errors, such as viewing a disease as if it is an adaptation. We assessed the quantity of errors in the most widely cited articles on evolutionary psychiatry and its relationship to citation frequency

**Methods:** Two reviewers searched PubMed, Web of Science, and Google Scholar on September 8, 2023, using specific search terms related to “evolution” and “psychiatry”, in order to find the most highly cited articles in the field. Based on the work of Nesse, we developed a measure for assessing the number of errors and overall quality in evolutionary psychiatry articles. We applied the measure to the 20 most highly cited articles, and calculated the correlations between article quality and number of errors with number of citations.

**Results:** Twenty highly cited articles, with a mean citation count of 758.95 and publication year range from 1964 to 2011, were rated. While the most highly cited articles had good quality on average, they also made important errors. There was no significant correlation of article quality or article errors and citation count.

**Conclusion:** Highly cited articles in evolutionary psychiatry demonstrated strengths but also exhibited weaknesses. The lack of a relationship of quality and error scores with citation rates suggests that other factors influence such citations. Future research should focus on achieving consensus on how best to assess the quality of evolutionary psychiatry articles and on what errors should be avoided.

## INTRODUCTION

Nesse and Williams ‘seminal text, “Why We Get Sick” argued that evolutionary theory is useful in understanding traits that leave members of a species vulnerable to disorders (Nesse and Williams, 2012). Evolutionary medicine and evolutionary psychiatry subsequently emerged as new disciplines, with specialized conferences, textbooks, and journals (Natterson-Horowitz et al., 2023). In a recent authoritative review of evolutionary psychiatry, Nesse outlined a range of possible explanations for vulnerability to mental disorders including mismatch with current environments and tradeoffs whose benefits come at the cost of decreased robustness (Nesse, 2023). Such explanations complement proximal biological explanations that describe mechanisms with more distal evolutionary ones that describe the origins of designs vulnerable to failure (Nesse, 2023).

While many articles on evolutionary psychiatry are strong, some make fundamental conceptual errors. Nesse has emphasized what features of an evolutionary medicine article make for a high quality publications, as well as a number of conceptual errors that seem to bedevil the field (Nesse, 2007). Good articles define their questions with precision and consider multiple possible explanations of observed phenomena. Errors in evolutionary medicine include viewing disorders as adaptations, and providing explanations based on what is good for the species (Nesse, 2007).

We developed a measure of the quality of and errors in evolutionary psychiatry articles, based on Nesse’s work (Nesse, 2007), and applied this to the most widely cited articles in the field. In this way, we aimed to investigate the quality of work that has been particularly influential, and to determine empirically whether the most highly cited articles were those with particularly high quality.

## METHODS

### Selection of Studies

Two reviewers (CB and CR) searched Pubmed, Web of Science and Google Scholar on8 September 2023 for relevant articles using search terms curated for each database, covering “evolution” and “psychiatry”. Two authors (CB and CR) reviewed the titles and abstracts of articles obtained by this search strategy. Included in this review were all articles that made evolutionary claims about particular mental disorders, excluding review articles and those focused on non-human research. The 20 most highly cited studies were included in this study.

### Rating of Studies

Articles were assessed for quality based on Nesse’s “How to test an evolutionary hypothesis about disease” (Table A – Section A) (Nesse, 2007), which lists 4 main objectives, these are: 1 - Define the object of explanation with great specificity; 2 - Specify all possible alternative hypotheses for why the trait is apparently suboptimal; 3 - Make explicit predictions from each hypothesis; 4. - Use all available evidence to test the predictions from all alternative hypotheses to arrive at a judgment about the contributions of different factors. Each objective has various provisos which are detailed in Table A.

To generate a quality score, each article was rated on each of the four objectives using a Likert scale ranging from 1 to 5: to what extent do you believe this paper adequately addresses the following, 1 – Strongly disagree; 2 – Disagree; 3 – Neither agree nor disagree (uncertain); 4 – Agree; 5 – Strongly agree. The four scores were summed to give a global quality score of 4 to 20. This score was then divided by 4 and assessed as follows 1-1.8 = Very poor; 1.8-2.6 = Poor; 2.6 – 3.4 = Average; 3.4-4.2 = Good; 4.2-5.0 = Very good.

Articles were then assessed for errors based on Nesse’s “Some common mistakes in testing evolutionary hypotheses about disease”(Nesse, 2007) (Table A – Section B). This table lists 10 possible errors, which are paraphrased here: 1 - Attempting to explain a disease as if it is an adaptation; 2 - Proposing an explanation based on what is good for the species; 3 - Proposing adaptive functions for rare genetic conditions; 4 - Confusing proximate and evolutionary explanations; 5 - Thinking that evidence for learning influencing a trait indicates that no evolutionary explanation is needed; 6 - Thinking that evidence for environmental or cultural differences in a trait is evidence against evolutionary influences; 7 - Confusing genetic explanations, especially behavioural genetic explanations, with evolutionary explanations; 8 - Failing to consider all of the alternative hypotheses; 9 - Assuming that evidence for one hypothesis is evidence against another; 10 - Presenting all the evidence in favour of a pet hypothesis and all the evidence against other hypotheses, instead of offering a balanced consideration of all evidence for and against all hypotheses.

We assessed each article for evidence of whether each of these errors was made. As points 1-4 above seem more serious errors than points 5-10, errors from points 1-4 were each given an error score of 1, while errors from points 5-10 were each given an error score of 0.5. Thus the total possible error score could range from 2 to 7. Finally, each article was assigned a global rating that ranged from 1 (Poor) to 5 (Excellent) (Table B).

All articles were scored independently by 2 reviewers (CB, CR). Differences were resolved by discussion or by bringing in a third reviewer (DJS).

### Statistical Analysis

Quality, error frequency, and overall impression scores were subjected to Cohen’s Kappa to assess inter-rater reliability (IRR), prior to resolution by discussion or the third reviewer (DJS). These results were interpreted as follows: values ≤ 0 as indicating no agreement and 0.01–0.20 as none to slight, 0.21–0.40 as fair, 0.41– 0.60 as moderate, 0.61–0.80 as substantial, and 0.81–1.00 as almost perfect agreement. Quality, error, and overall impression scores for each article were then correlated with citation counts and significance was assess using a two-tailed T-test. Statistical analysis was performed using IBM SPSS Statistics (version 26.0, IBM, USA) were used for analysis and differences were considered significant where p<0.05.

## RESULTS

### Included studies

We analyzed the 20 most highly cited articles in evolutionary psychiatry (Allen and Badcock, 2003, Andrews and Thomson, 2009, Bateson et al., 2011, Brüne, 2005, Crespi and Badcock, 2008, Crow, 2000, Gilbert and Allan, 1998, Hagen, 1999, Huxley et al., 1964, Jonason et al., 2009, Klein, 1993, Marks and Nesse, 1994, Nesse, 1998, Nesse, 2000, Nesse, 2001, Mealey, 1995, Nettle and Clegg, 2006, Price et al., 1994, Sloman et al., 2003, Watson and Andrews, 2002). The lowest cited article had 310 citations, while the highest cited article had 1546 citations, with a mean of 758.95 and median of 627 citations. The earliest published article was from 1964, with the most recent article being from 2011, with a mean publication year of 2000 and a standard deviation of 10.01. Table C shows the included articles, with study ID and citation number.

### Study Ratings

Table D shows the results of the quality scores, with included means and standard deviations. The overall quality score showed a mean of 3.6, standard deviation of 0.6. This falls on the low end of the good score of 3.4-4.2. Notably the highest scoring section was F1 (M = 3.9, SD = 0.6). There were varying degrees of IRR, with F4 showing the highest level (k = 0.4).

Table E shows the results of the error scoring, with total number of times the error was made and the percentage. Notably, FN8 and FN10 were found to be the most common, each being made by ten of the articles assessed (50%). FN5 and FN6 were not found to be made in the assessed articles. The item with the highest IRR was FN1 (k = 0.857).

The mean general impression score was 3.0, with a standard deviation of 1.5. There was moderate agreement in terms of IRR (k = 0.6).

### Statistical Analysis

There was no significant correlation between quality score and citation number (r(18) = −.06, p = ns), or publication year (r(18) = .22, p = 0.343). There was no significant correlation between error score and citation number (r(18) = .28, p = 0.234), or publication year (r(18) = −.32, p = 0.174). There was no significant correlation between general impression score and citation number (r(18) = −.09, p = 0.704) or publication year (r(18) = .14, p = 0.562). There was a significant correlation between quality score and general impression score (r18) = 0.728, p < 0.001), and there was a significant correlation between higher quality score and lower error score (r(18) = 0.62, p = 0.03). Specific correlations between individual quality and error scores are reported in Tables F1 and F2.

## DISCUSSION

The main findings of this research were 1) the most highly cited articles have good quality on average, 2) the most highly cited articles make important errors, 3) there was no significant relationship between quality of article and number of citations.

Overall, the top 20 mostly highly cited articles achieved a mean on the low end of the “good” rating for each of the 4 positive scoring metrics (3.6-3.9; with good considered 3.4-4.2), with the overall positive score also achieving a “good” rating with 3.6, std 0.6. The included articles scored a mean of 3.9 on F1 (Define the object of explanation with great specificity), and a mean of 3.6 on F3 (Make explicit predictions from each possible hypothesis), indicating that these areas represent strengths of the included articles. Conversely, F2 (Specify all possible alternative hypotheses for why the trait is apparently suboptimal) and F4 (Use all available evidence to test the predictions from all alternative hypotheses to arrive at a judgment about the contributions of different factors. had the lowest mean score of included articles (3.6), indicating that these are areas that future articles can improve upon.

Of the errors made, the most common were FN8 (Failing to consider all of the alternative hypotheses) and FN10 (Presenting all the evidence in favour of a pet hypothesis and all the evidence against other hypotheses, instead of offering a balanced consideration of all evidence for and against all hypotheses). 50% of the assessed articles made one or both of these errors. As F2 and FN8 cover similar ideas, their strong correlations are understandable; as are the correlations between F3, F4 and FN10. The high number of correlations of FN8 and FN10 with a range of other items suggest that these are decisive aspects of the overall assessment, and deserve particular attention in future work.

None of the top 20 articles most highly cited made FN5 (Thinking that evidence for learning influencing a trait indicates that no evolutionary explanation is needed) or FN6 (Thinking that evidence for environmental or cultural differences in a trait is evidence against evolutionary influences) as an error. These errors may, however, be more prevalent in less cited evolutionary psychiatry articles. A key limitation of the current project is that we cannot generalize from the work here to the field of evolutionary psychiatry as a whole. Furthermore, we acknowledge that different databases provide different citation rates; here we relied on Google Scholar.

Quality ratings and number of errors did not influence citation rates; possible explanations are beyond the scope of this review. Previous work suggests that in many fields quality and methodological rigor are not necessarily correlated with citation number (Lopez et al., 2017, Mackinnon et al., 2018). In neuroscience, orthopaedics and plastic surgery citation numbers are influenced by factors including age of the article, study design, level of evidence, conflict of interest disclosures, and number of authors (Jamjoom et al., 2022, Lopez et al., 2017, Okike et al., 2011).

In conclusion, our results suggest that the most highly cited articles in evolutionary psychiatry have both strengths and weaknesses. Strengths include defining the research question with precision and predicting the answers based on a specific hypothesis. Weaknesses include failing to consider and specify alternate hypotheses, and presenting evidence in favour of a pet hypothesis while not using all available evidence. The lack of a relationship of quality and error scores with citation rates suggests that other factors influence such citations. While the field has significant promise, further work is needed to achieve consensus on how best to assess the quality of evolutionary psychiatry articles and on what errors should be avoided.

## Supporting information

Table A - Nesse criteria

Table B - FGI scoring

Table C - Included articles

Table D - Positive Score

Table E - Negative score

## Data Availability

All data produced in the present work are contained in the manuscript

