## Supplementary material for "HIGHLY CITED ARTICLES IN EVOLUTIONARY PSYCHIATRY: ASSESSMENT WITH A QUALITY AND ERROR RATING SCALE": Table A - Nesse criteria

### Table A - How to test an evolutionary hypothesis about disease – adapted from Randolph Nesse

**SECTION A**

**F1. Define the object of explanation with great specificity.**

a. A trait shaped by natural selection wiIl usuaIly be the object of explanation.
b. A disease is an appropriate object only if the hypothesis is that the "disease" is actuaIly a defense, or otherwise increases fitness.
c. Usually the object is a trait that is universal in the species and that makes an organism vulnerable to a disease.
d. If the object is not a universal trait, justify the exception.

i. The most common exceptions are traits that reflect genetic differences in subpopulations responding to local environment, e.g., sickle cell aIleles that protect against malaria.
ii. Other exceptions are facultative adaptations that explain individual differences, e.g., the number of sweat glands increases as a function of early exposure to high temperatures.
iii. If the trait is a behavior, do not explain the behavior, instead explain how selection shaped the behavior regulation system.

e. Do not propose an evolutionary explanation for why one individual gets a disease and another does not. Evolutionary explanations are about populations. They may however, predict who get a disease.
f. Evolutionary explanations are not alternatives to proximate explanations. Evidence about proximate mechanisms is often useful in assessing a hypothesis, however, especially when the hypothesis is that vulnerability to disease results from constraints or trade-offs.

**F2. Specify all possible alternative hypotheses for why the trait is apparently suboptimal. There are six main possibilities:**

a. The environment has changed faster than selection and the disease results from this mismatch.
b. The relevant environmental factor is a pathogen that evolves faster than host defenses.
c. Constraints, e.g., natural selection's limited ability to clear mutations or correct a fundamentally defective design, leave the organism vulnerable.
d. The trait offers trade-off compensatory benefits that account for apparently suboptimal features.
e. The trait offers benefits to reproduction or to kin that are greater than the costs to the individual.
f. The trait is not a disease at all, but a useful protective response such as pain or fever.

**F3. Make explicit predictions from each possible hypothesis.**

a. If relevant data from other species can be obtained and analyzed, use the comparative method.
b. Otherwise, try to make predictions about aspects of the trait or its regulation, preferably previously unrecognized and quantitative.
c. Other useful predictions may be made about the relative fitness of individuals with and without the trait in different environments.
d. Predictions about proximate aspects of the trait may be possible.

**F4. Use all available evidence to test the predictions from all alternative hypotheses to arrive at a judgment about the contributions of different factors.**

a. Note that multiple factors often operate together to an apparently suboptimal trait. This is quite different from proximate explanations where evidence for one alternative usually weighs against others.
b. Many hypotheses can be falsified because they are inconsistent with evolutionary theory.
c. Others can be falsified by experiments that show that a trait does not serve the proposed function.
d. Assess the overall plausibility of the proposal and the relative viability of alternative hypotheses.

**SECTION B**
**Some Common Mistakes in Testing Evolutionary Hypotheses About Disease**

The guidelines below tacitly describe a variety of possible errors, some of which are made explicit below.

**FN1**. Attempting to explain a disease: Instead, reformulate the question as an explanation for vulnerability to a disease.

**FN2**. Proposing an explanation based on what is good for the species: This is group selection, an elementary error. Almost all evolutionary explanations must be based on advantages to genes or individuals.

**FN3**. Proposing adaptive functions for rare genetic conditions: There are sometimes evolutionary reasons why deleterious mutations stay in the gene pool, but the explanation is hardly ever some useful function of the disease itself.

**FN4**. Confusing proximate and evolutionary explanations: This is a common and serious mistake. Knowledge about how the body works can be very useful in assessing an evolutionary hypothesis, but it is no substitute for an evolutionary explanation.

**FN5**. Thinking that evidence for learning influencing a trait indicates that no evolutionary explanation is needed: Learning is a capacity shaped by natural selection, and the pathologies that arise from learning mechanisms, such as phobias, are likely to harm fitness.

**FN6**. Thinking that evidence for environmental or cultural differences in a trait is evidence against evolutionary influences: Natural selection shaped the behavioral mechanisms that give rise to culture, and environments and culture influence human behavior and fitness strongly. An evolutionary approach to behavior does not imply that behavior is somehow "determined by the genes," only that the mechanisms that give rise to behavior and culture were shaped by natural selection. These mechanisms obviously are capable of profound flexibility, with attendant major benefits and costs.

**FN7**. Confusing genetic explanations, especially behavioral genetic explanations, with evolutionary explanations: Traits need evolutionary explanations whether or not individual variations arise from genetic variations.

**FN8**. Failing to consider all of the alternative hypotheses: This is very common and very serious. All too often an author will propose one possibility without making the alternatives explicit

**FN9**. Assuming that evidence for one hypothesis is evidence against another: Multiple factors may all contribute to a complete explanation and they may interact in complex ways. Correct explanations often incorporate multiple explanatory factors.

**FN10**. Presenting all the evidence in favor of a pet hypothesis and all of the evidence against other hypotheses, instead of offering a balanced consideration of all evidence for and against all hypotheses: This is rhetoric, not science. It is observed commonly, for good reasons arising from human nature, not just in testing evolutionary hypotheses but across the range of sciences. Nonetheless, such advocacy should be avoided.

Final Global Impression composing aspects of both the positive and negative scores (rating that ranged from 1 (Poor) to 5 (Excellent)
