## Supplementary material for "HIGHLY CITED ARTICLES IN EVOLUTIONARY PSYCHIATRY: ASSESSMENT WITH A QUALITY AND ERROR RATING SCALE": Table B - FGI scoring

**Table B – Final Global Impression (GGI) scoring**

| | | |
|---|---|---|
| 1 | Poor | Makes an elementary mistake about evolutionary theory (attempts to explain a disease, proposes an explanation based on what is good for the species, proposes adaptive functions for rare genetic conditions, confuses proximate and evolutionary explanations). |
| 2 | Problematic | Makes no elementary mistakes about evolutionary theory, but does not represent the best of evolutionary theory (focused on proximal explanations rather than evolutionary ones). |
| 3 | Middling | Is consistent with good evolutionary theory, but defines the object of explanation with limited specificity, with limited discussion of alternate hypotheses or explicit predictions, or without clearly demonstrating clinical implications. |
| 4 | Good | Defines the object of explanation with some specificity, and has some discussion of alternative hypothesis or explicit predictions, and has clear discussion of the clinical implications. |
| 5 | Excellent | Defines the object of explanation with great specificity (a trait shaped by natural selection, that makes an organism vulnerable to disease), specifies alternate hypotheses about why the trait is suboptimal, make explicit predictions from the hypotheses, and if true promises to change clinical practice. |

Final Global Impression composing aspects of both the positive and negative scores (rating that ranged from 1 (Poor) to 5 (Excellent)
