## Supplementary material for "HIGHLY CITED ARTICLES IN EVOLUTIONARY PSYCHIATRY: ASSESSMENT WITH A QUALITY AND ERROR RATING SCALE": Table C - Included articles

| Article ID | Citations | Pub. Year |
|---|---|---|
| Allen2003 | 340 | 2003 |
| Andrews2009 | 382 | 2009 |
| Bateson2011 | 146 | 2011 |
| Brune2005 | 850 | 2005 |
| Crespi2008 | 435 | 2008 |
| Crow2000 | 307 | 2000 |
| Gilbert1998 | 655 | 1998 |
| Hagen1999 | 183 | 1999 |
| Huxley1964 | 188 | 1964 |
| Jonason2009 | 626 | 2009 |
| Klein1993 | 1000 | 1993 |
| Marks1994 | 448 | 1994 |
| Mealey1995 | 676 | 1995 |
| Nesse1998 | 130 | 1998 |
| Nesse2000 | 625 | 2000 |
| Nesse2005 | 309 | 2005 |
| Nettle2006 | 204 | 2006 |
| Price1994 | 405 | 1994 |
| Sloman2003 | 170 | 2003 |
| Watson2002 | 187 | 2002 |

Table C shows the included articles, with study ID and citation number as per Scopus (Current as of 07/01/2025)
