## Supplementary material for "HIGHLY CITED ARTICLES IN EVOLUTIONARY PSYCHIATRY: ASSESSMENT WITH A QUALITY AND ERROR RATING SCALE": Table D - Positive Score

**Table D – Positive scores**

| | F1 | F2 | F3 | F4 | FP |
|---|---|---|---|---|---|
| | 3 | 2 | 3 | 3 | 2.75 |
| | 3 | 3 | 3 | 2 | 2.75 |
| | 3 | 3 | 3 | 3 | 3.00 |
| | 3 | 3 | 3 | 3 | 3.00 |
| | 4 | 2 | 4 | 3 | 3.25 |
| | 3 | 4 | 3 | 3 | 3.25 |
| | 4 | 3 | 3 | 3 | 3.25 |
| | 4 | 3 | 3 | 3 | 3.25 |
| | 4 | 3 | 3 | 4 | 3.5 |
| | 4 | 3 | 4 | 4 | 3.75 |
| | 4 | 4 | 4 | 3 | 3.75 |
| | 4 | 4 | 3 | 4 | 3.75 |
| | 4 | 4 | 4 | 4 | 4.00 |
| | 4 | 4 | 4 | 4 | 4.00 |
| | 4 | 4 | 4 | 4 | 4.00 |
| | 4 | 4 | 4 | 4 | 4.00 |
| | 5 | 4 | 4 | 4 | 4.25 |
| | 5 | 4 | 4 | 4 | 4.25 |
| | 4 | 5 | 4 | 4 | 4.25 |
| | 4 | 5 | 5 | 5 | 4.75 |
| Mean | M = 3.85 | M= 3.55 | M= 3,60 | M= 3.55 | M= 3.64 |
| SD | SD = 0.59 | SD= 0.83 | SD= 0,60 | SD= 0.69 | SD= 0.56 |

Table D shows the results of the quality scores ordered by total score, with included means and standard deviations
