## Supplementary material for "HIGHLY CITED ARTICLES IN EVOLUTIONARY PSYCHIATRY: ASSESSMENT WITH A QUALITY AND ERROR RATING SCALE": Table E - Negative score

**Table E – Negative scores**

| | FN1 | FN2 | FN3 | FN4 | FN5 | FN6 | FN7 | FN8 | FN9 | FN10 | FTN |
|---|---|---|---|---|---|---|---|---|---|---|---|
| | 0 | 0 | 0 | 0 | 0 | 0 | 0 | 0 | 0 | 0 | 0 |
| | 0 | 0 | 0 | 0 | 0 | 0 | 0 | 0 | 0 | 0 | 0 |
| | 0 | 0 | 0 | 0 | 0 | 0 | 0 | 0 | 0 | 0 | 0 |
| | 0 | 0 | 0 | 0 | 0 | 0 | 0 | 0 | 0 | 0 | 0 |
| | 0 | 0 | 0 | 0 | 0 | 0 | 0 | 0 | 0 | 0 | 0 |
| | 0 | 0 | 0 | 0 | 0 | 0 | 0 | 0 | 0 | 0 | 0 |
| | 0 | 0 | 0 | 0 | 0 | 0 | 0 | 0 | 0 | 0 | 0 |
| | 0 | 0 | 0 | 0 | 0 | 0 | 0 | 0 | 0 | 0 | 0 |
| | 0 | 0 | 0 | 0 | 0 | 0 | 0 | 1 | 0 | 1 | 1 |
| | 0 | 0 | 0 | 0 | 0 | 0 | 0 | 1 | 0 | 1 | 1 |
| | 1 | 0 | 0 | 0 | 0 | 0 | 0 | 1 | 0 | 1 | 2 |
| | 0 | 0 | 0 | 0 | 0 | 0 | 1 | 1 | 1 | 1 | 2 |
| | 1 | 0 | 1 | 0 | 0 | 0 | 0 | 1 | 0 | 1 | 3 |
| | 1 | 0 | 1 | 1 | 0 | 0 | 0 | 1 | 0 | 1 | 4 |
| | 0 | 0 | 0 | 0 | 0 | 0 | 0 | 1 | 0 | 0 | 0.5 |
| | 0 | 0 | 0 | 0 | 0 | 0 | 0 | 0 | 0 | 1 | 0.5 |
| | 0 | 0 | 0 | 0 | 0 | 0 | 0 | 1 | 1 | 1 | 1.5 |
| | 1 | 0 | 0 | 0 | 0 | 0 | 0 | 0 | 1 | 0 | 1.5 |
| | 0 | 0 | 0 | 1 | 0 | 0 | 1 | 1 | 0 | 1 | 2.5 |
| | 0 | 1 | 1 | 0 | 0 | 0 | 0 | 1 | 1 | 1 | 3.5 |
| Times made | 4 | 1 | 3 | 2 | 0 | 0 | 2 | 10 | 4 | 10 | |
| % articles | 20% | 5% | 15% | 10% | 0% | 0% | 10% | 50% | 20% | 50% | |

Table E shows the results of the error scoring, ordered by total score, with total number of times the error was made and the percentage.
